# Systematic review and individual patient data meta-analysis on glucose- 6 – phosphate dehydrogenase activities measured by a semi-quantitative handheld Biosensor

**DOI:** 10.1101/2024.12.20.24319407

**Authors:** Benedikt Ley, Laura Rojas Vasquez, Avyinaeesh Sitsabasan, Bipin Adhikari, Nabaraj Adhikari, Mohammad Shafiul Alam, Santasabuj Das, Prakash Ghimire, Marcus V. G. Lacerda, Ric N. Price, Komal Raj Rijal, Lorenz von Seidlein, Arunansu Talukdar, Kamala Thriemer, Ari Winasti Satyagraha, Arkasha Sadhewa, Megha Rajasekhar, Robert J. Commons

## Abstract

Measurement of glucose-6-phosphate dehydrogenase (G6PD) activity guides hypnozoitocidal treatment of *P. vivax* malaria. The G6PD Standard (SDBiosensor, Republic of Korea) here referred to as “Biosensor” is a quantitative point-of-care diagnostic that measures G6PD activity in U/gHb . The manufacturer recommends cutoffs to define G6PD deficient (≤4.0U/gHb), intermediate (4.1-≤6.0U/gHb) and normal (>6.0U/gHb) individuals. The aim of this individual patient data (IPD) meta-analysis was to evaluate these cutoffs (CRD42023406595).

A systematic review identified studies reporting population-level G6PD activity measured by Biosensor, published between January 2017 and May 2023. IPD were collated and standardised. The adjusted male median (AMM) was defined as 100% activity and calculated across all studies (universal AMM) and separately for each setting. The proportion of participants classified as deficient or intermediate were compared using the manufacturer-recommended cutoffs and 30% and 70% of the universal AMM and setting-specific AMM. Associations between G6PD activity and blood sampling method, malaria status, and age were assessed.

Eleven studies with 9,724 participants from eight countries were included in this analysis. The universal AMM was 7.7U/gHb and the setting-specific AMMs ranged from 6.2U/gHb to 9.9U/gHb. When using the universal AMM, 4.2% of participants were classified as deficient and 11.9% as intermediate or deficient. The corresponding values were 3.9% and 10.8% for setting-specific cutoffs, and 7.2% and 18.3% for manufacturer-recommended definitions for deficients and intermediates respectively. The manufacturer-recommended cutoff for deficient individuals fitted the distribution of G6PD activities better than definitions based on the percentage of AMM. There was no significant association between malaria status or blood sampling method and G6PD activity. Measured G6PD activity decreased in children 1 to 5 years and plateaued thereafter.

The manufacturer’s recommended cutoff is conservative but more reliable at categorising G6PD deficient individuals than those based on calculations of 30% activity using the AMM. The observed decrease in G6PD activity in children between 1 to 5 years of age warrants further investigation.

## Background

Nearly 3.3 billion people are at risk of a *Plasmodium vivax* (*P. vivax*) infection [1–3]. In contrast to other malaria species *P. vivax* forms dormant liver stage hypnozoites that relapse weeks to months after a primary infection causing significant morbidity and mortality [4]. These hypnozoites represent a silent reservoir that complicates the elimination of *P. vivax* [5]. The 8-aminoquinolines, primaquine and tafenoquine, are the only licensed drugs that kill hypnozoites; although both drugs can cause severe haemolysis in patients with low glucose-6-phosphate dehydrogenase enzyme (G6PD) activity [6, 7].

G6PD is found in all eukaryotic cells and is essential in human red blood cells (RBCs) to maintain their redox potential [8, 9]. Enzyme activity is measured in international units (U), normalised by a haemoglobin (Hb) measurement (U/gHb) taken at the same time [10]. Since the phenotypic reference method, spectrophotometry, is poorly standardised, the output in U/gHb is converted to percent activity (%) based on the adjusted male median (AMM), to enable comparison across populations [11]. To calculate the AMM, the median activity of all male individuals from the same population is calculated, all observations with values less than 10% of this median are then excluded and the median activity is recalculated from the remaining subset and defined as 100% activity [12]. More than 230 clinically relevant genetic G6PD variants have been described to date, associated with reduced G6PD activities, collectively called G6PD deficiency [8, 13, 14].

The G6PD gene is located on the X-chromosome. Males have one copy of the gene and are hemizygous deficient or normal. Females have two copies and can be either homozygous deficient, homozygous normal, or heterozygous for the gene. Most hemizygous and homozygous deficient individuals have phenotypic G6PD activities below 30% and G6PD normal individuals have activities above 70% to 80% [15]. In comparison, heterozygous females have a wide range of G6PD activities from deficient to normal [8]. Accordingly, G6PD activities within a population show a bimodal distribution, with activities of the majority of individuals normally distributed around the 100% activity mark and a second smaller peak representing hemizygous and homozygous deficient individuals (Fig 1) [16].

**Fig 1:**
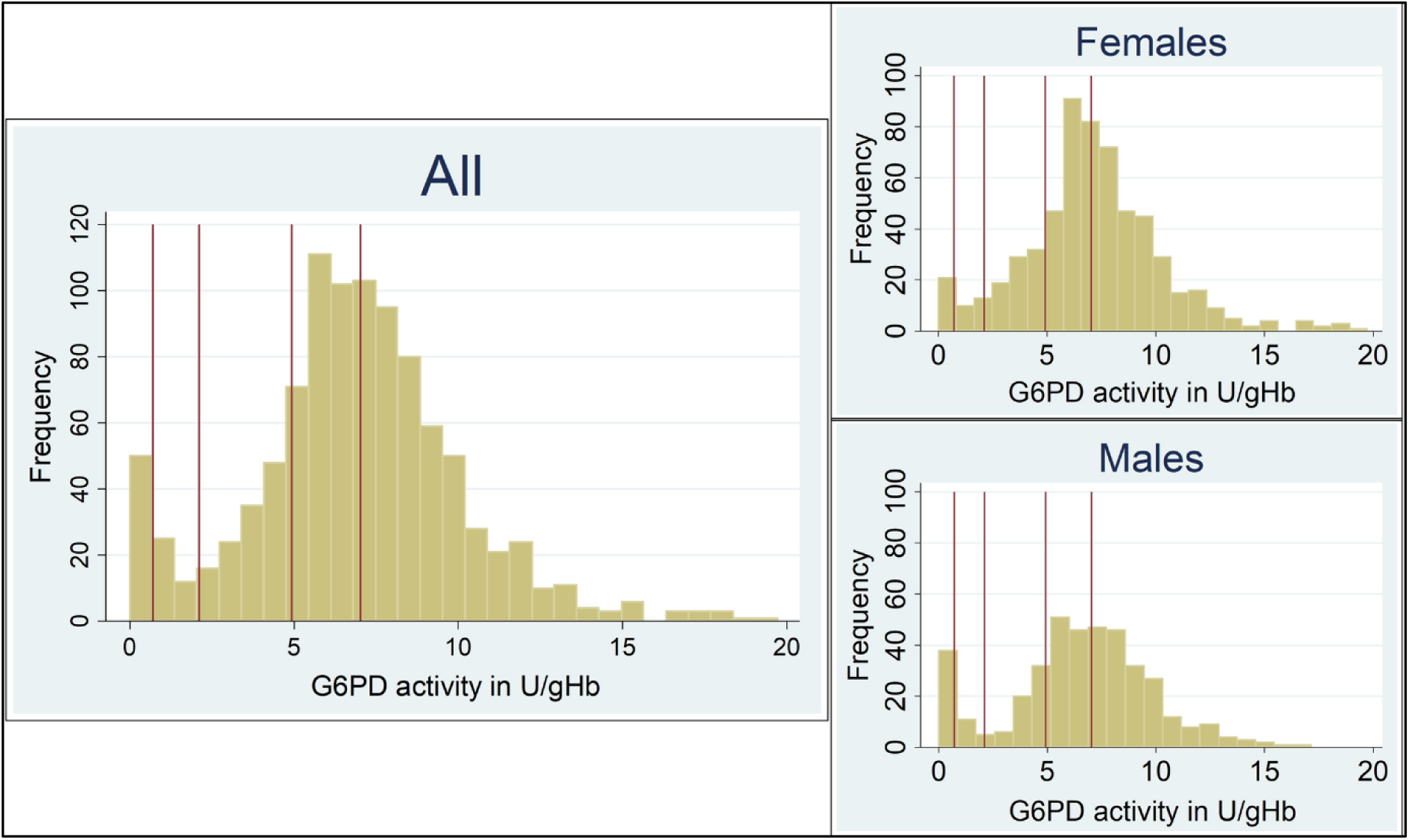
G6PD activities in samples collected from participants in Bangladesh during a cross-sectional survey measured by spectrophotometry *Vertical lines from left to right indicate 10%, 30% 70% and 100% G6PD activity* [16]

The World Health Organization (WHO) recommends testing patients for G6PD deficiency prior to treatment with daily primaquine regimens to identify patients with reduced activity and tailor their treatment accordingly [17]. Patients with G6PD activity <30% should not receive 7- or 14-day primaquine regimens. Tafenoquine is only recommended for patients with ≥70% G6PD activity [18, 19]. In 2022 the WHO revised their classification of G6PD variants, recommending that any variant with a median activity <45% (Class B) by spectrophotometry should be considered clinically significant [14, 20, 21]. Although spectrophotometry is the reference method to measure G6PD activity, it is unsuitable for point of care testing [22]. To date, the only point of care test that can reliably discriminate activities between 30% and 70% is the handheld semi-quantitative Biosensor from SD Biosensor (STANDARD G6PD, ROK; Biosensor) [22–25]. The Biosensor generates G6PD (in U/gHb) and haemoglobin (in g/dL) readings from 10 µl of capillary or venous blood within two minutes. The manufacturer recommends thresholds of 4.0 U/gHb and 6.0 U/gHb to discriminate deficient and intermediate activities respectively [26]. Alignment of these cutoffs with local AMM-derived 30% and 70% cutoffs have varied considerably [27]. This individual patient data pooled analysis aimed to assess whether the manufacturer-recommended cutoffs for the Biosensor are universally applicable.

## Methods

### Search strategy and selection criteria

A systematic review of Medline, Web of Science, Cochrane Central, Scopus, Clinicaltrials.gov and the WHO International clinical trials registry platform was undertaken to identify studies that measured population-based G6PD activity using the G6PD Standard point of care test (Biosensor) manufactured by SD Biosensor (Republic of Korea) between January 1, 2017, and May 17, 2023, in any language. Search terms used were ((G6PD) OR (G6PD deficiency) OR (Glucose-6-phosphate dehydrogenase) OR (Glucose-6-phosphate dehydrogenase deficiency)) AND ((G6PD Standard) OR (SDBiosensor) OR (SD Biosensor) OR (Standard G6PD) OR tafenoquine). Studies were included if they reported activities of at least 36 randomly selected male individuals older than 1 year from a population expected to have a G6PD distribution consistent with the local population. Four reviewers undertook the systematic review (AS, BL, LRV and RJC) with each article screened by two reviewers and discrepancies resolved by discussion. The protocol was registered with PROSPERO CRD42023406595.

Investigators of studies eligible for inclusion were invited to contribute individual patient data in addition to any data from eligible but unpublished studies. Data collected included age, sex, malaria status, G6PD activity (in U/gHb) and haemoglobin (in g/dL) measured by Biosensor, date of G6PD measurement, methodology of blood collection (capillary versus venous), pregnancy status, date of last malaria episode, and history of giving birth, blood loss or major surgery in the previous 6 months. Individual patient data were excluded if missing age, sex, G6PD activity, or haemoglobin, or if haemoglobin was <7 g/dL on the Biosensor measurement, as recommended by the manufacturer.

Data were obtained according to ethical approvals from the original study. Our analysis did not require additional ethical approval as the original data approved by ethical review boards were anonymised and unable to be linked to individuals.

### Definitions

Each combination of country and study group was defined as a ‘setting’. The blood sampling technique was categorised as venous or capillary. The AMM was defined as 100% activity and calculated from male participants above one year of age who tested negative for malaria by microscopy or rapid diagnostic test (RDT) and after removal of participants with the lowest 10% of reported G6PD activities [12]. The AMM was calculated for the entire data set (universal AMM) and separately for each setting (setting-specific AMM). G6PD deficient and intermediate status of all included participants were categorised repeatedly: i) as measured activity of less than 30% and 70% of the universal AMM, ii) as measured activity of less than 30% and 70% of the setting-specific AMM, and iii) when applying the manufacturer recommended cut-off of 4.0 U/gHb and 6.0 U/gHb, respectively.

### Statistical Analysis

All observations were categorised three times based on the different definitions of deficient and intermediate activities and proportions of deficient and intermediate individuals were compared. The distribution of G6PD activities was plotted by sex for the overall population and by setting. The association between blood sampling method (capillary or venous) and G6PD activity was assessed in studies that sampled using both capillary and venous methods. The relationship was analysed using multivariable linear regression adjusting for age, sex, baseline Hb, and malaria status with robust errors and clustering by setting. Using a similar multivariable linear regression model restricted to studies which included participants both with or without malaria, the association between malaria presence and G6PD activity was investigated. The relationship between age and G6PD activity was also investigated in a similar model. Due to a non-linear relationship between age and G6PD activity, a post hoc analysis was undertaken with a generalised estimating equations model to visualise the relationship between G6PD activity and age with fractional polynomial terms fitted for age, adjustment for age, sex, baseline Hb, and blood sampling method and clustering by setting. This relationship was explored separately in all participants, in males and females. Separate sensitivity analyses were undertaken restricted to males with i) a haemoglobin >10 g/dL and ii) excluding those with an activity ≤4.0 U/g Hb (presumed deficient).

The within-study risk of bias assessment was performed using the QUADAS-2 tool [28]. Analyses were undertaken in Stata (version 17).

## Results

A total of 322 studies were found in the databases, published between January 1, 2017, and May 17, 2023, of which 300 were excluded after screening titles and abstracts (Figure 2). A further 18 studies were excluded because G6PD activity was not measured by Biosensor (n=12), participants were not selected randomly from the whole population (n=4) or the trial was ongoing (n=2). A total of four studies met the eligibility criteria [29–32] and data were available and shared by investigators. Data on sex were not recorded consistently across G6PD categories for one study leading to its exclusion. Data from eight studies unpublished as of May 17, 2023, were also available, providing a total of 11 studies on 10,905 participants [33, 34]. Data from 1,181 participants were excluded due to missing variables and haemoglobin measurements <7 g/dL, resulting in 9,724 participants from 11 studies included in the analysis .

**Figure 2.**
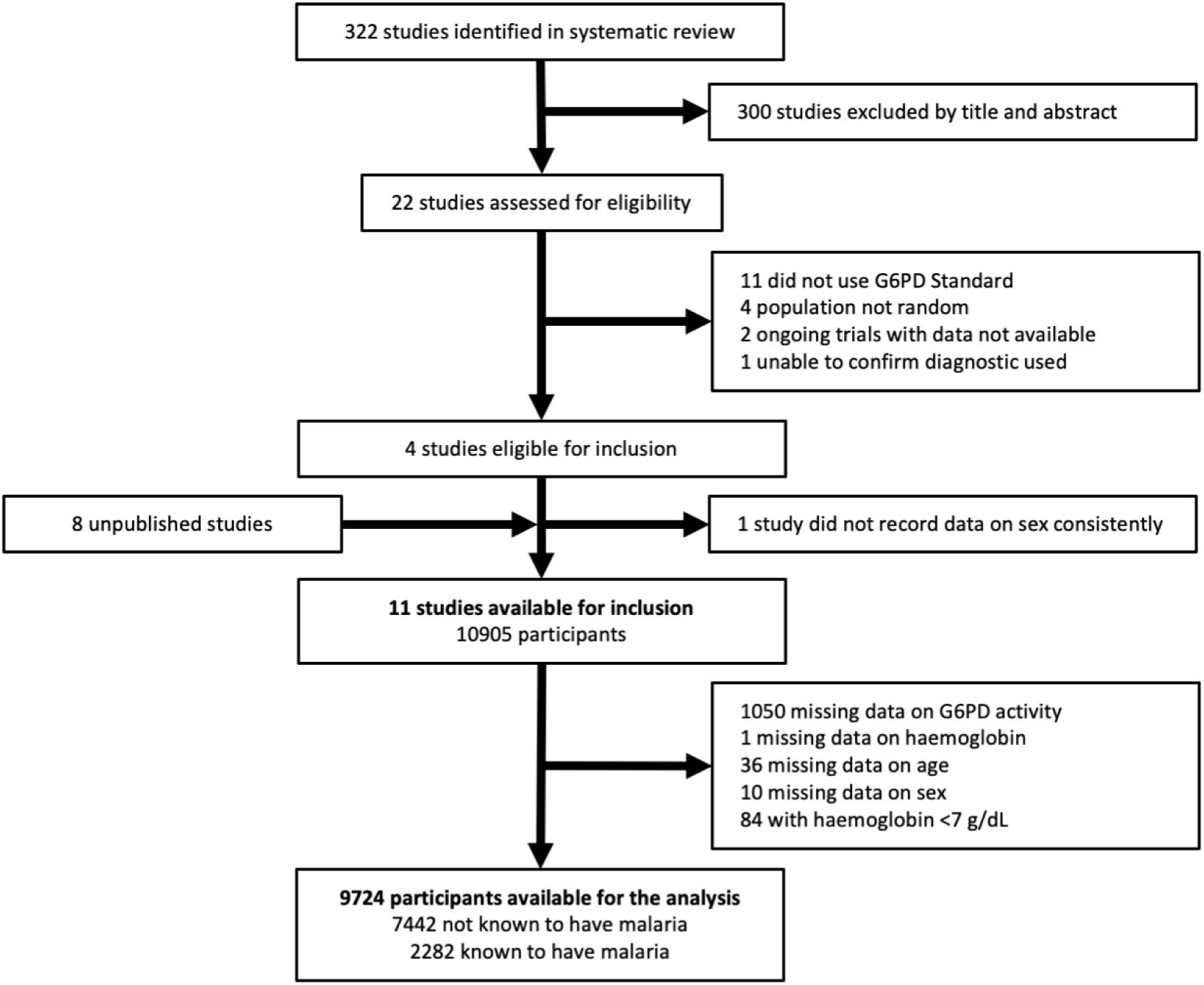
Flow diagram of patient inclusion

Data were included from 8 countries: Bangladesh, Brazil, Cambodia, Ethiopia, India, Indonesia, Nepal, and the United States of America. The median age was 30.0 years (interquartile range (IQR) 21.0-43.0), of whom 4,237 (43.6%) were female and 8,938 (91.9%) were sampled using capillary sampling methods. Malaria was recorded in 2,282 (23.5%) participants. No consistent data were available on pregnancy, previous malaria episodes, recent surgery, recent blood loss, or recent blood transfusions. There were 14 unique settings (country and study categories), of which 2 (14%) had unclear or possible risk of bias (Table S1 - QUADAS).

The distribution of G6PD activities in male participants showed two peaks consistent with presumed G6PD deficient and G6PD normal populations (Figure 3). Visually, the nadir in the overlapping region between these peaks occurred at ∼3.5 U/gHb. The universal AMM, derived from 3,959 males without parasitaemia, was 7.7 U/gHb and from this the 30% cutoff was determined to be 2.3 U/gHb and the 70% cutoff was determined to be 5.4 U/gHb (Table 2 and Figure 3). The AMM calculated for each setting varied substantially from 6.2 U/gHb in one study from Nepal to 9.9 U/gHb in a study in Bangladesh [33, 34]. The setting-specific cutoffs for 30% G6PD activity ranged from 1.9 U/g Hb to 3.0 U/g Hb and the corresponding setting-specific 70% cutoffs varied from 4.3 U/gHb to 6.9 U/gHb (Table 2).

**Figure 3.**
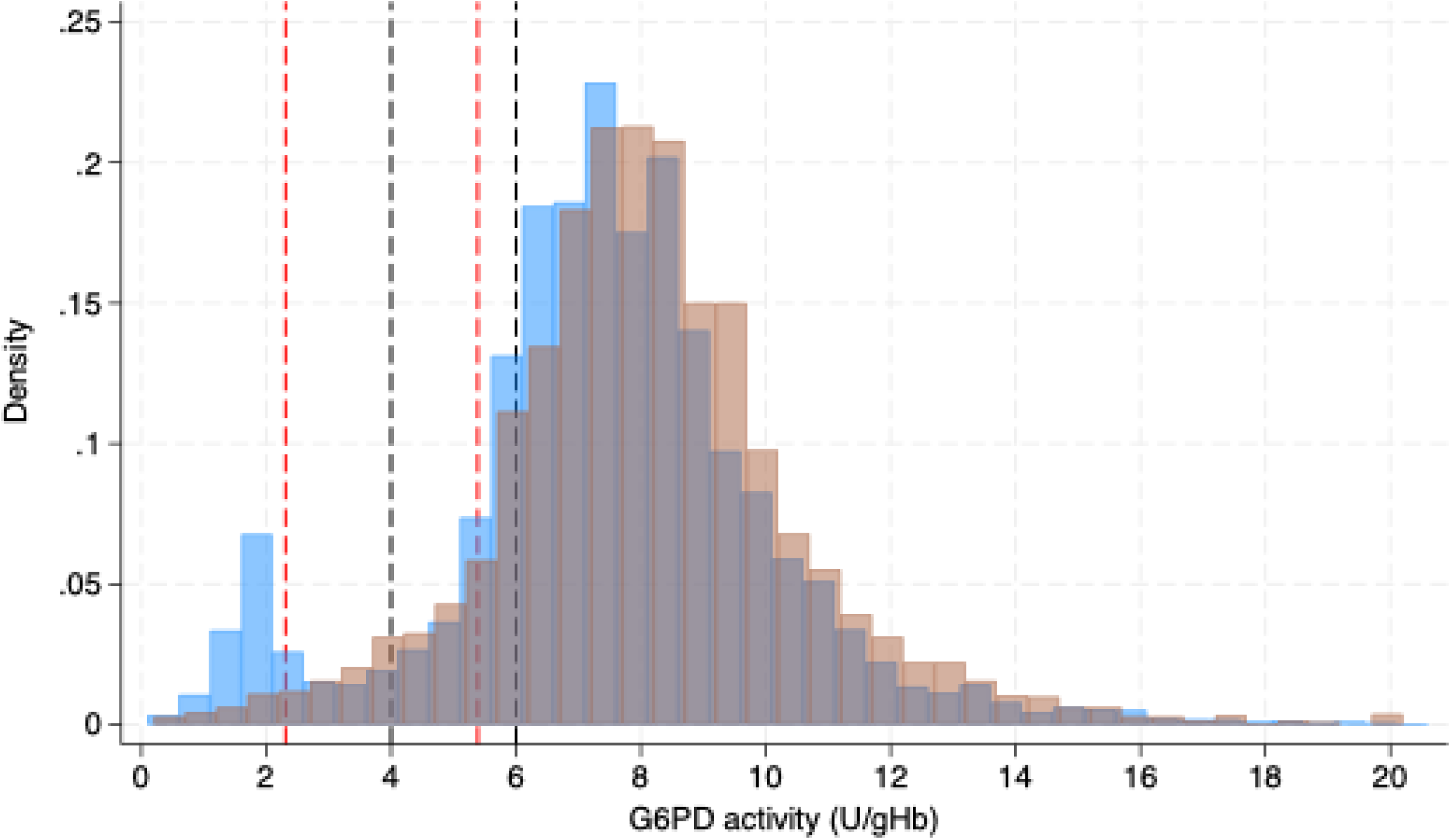
Density of G6PD activity by sex Blue bars correspond to males. Orange bars correspond to females. Red dotted lines correspond to 30% and 70% activity of the universal AMM. Black dotted lines correspond to the manufacturer recommended cutoff of 4.0 U/gHb and 6.0 U/gHb.

Using the manufacturer’s cutoff of 4.0 U/gHb, 7.2% (699) of all participants in this analysis were classified as deficient (72.8% (509) were males), but this decreased to 4.2% (411) when the universal 30% cutoff of 2.3 U/gHb was applied (Figure 3), and 3.9% (301) when the setting-specific 30% cutoffs were applied (Table 2 and Figure 4). Using the manufacturer’s cutoff of 6.0 U/gHb, 18.3% (1,777) of participants were classified as having intermediate or deficient G6PD activity (<70%) and this decreased to 11.9% (1,156) when the universal cutoff was applied and 10.8% (830) when the setting-specific cutoffs were applied. Of the 1,078 patients with intermediate activity (>4.0-6.0 U/gHb) using the manufacturer’s cutoffs, 60.1% (648) were male. The distribution of G6PD activities varied substantially between settings (Figure 4 and Tables 1-2).

**Figure 4.**
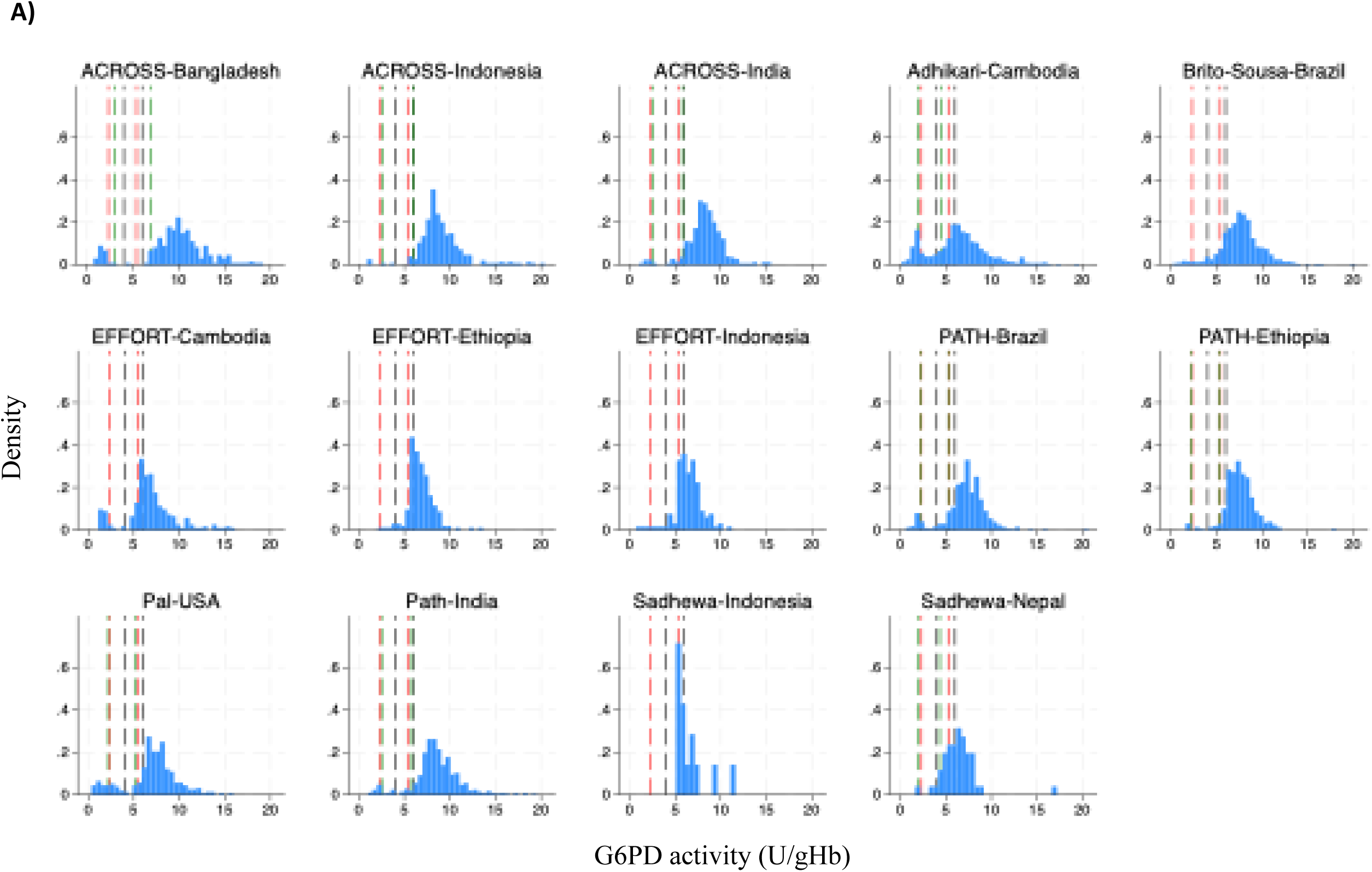
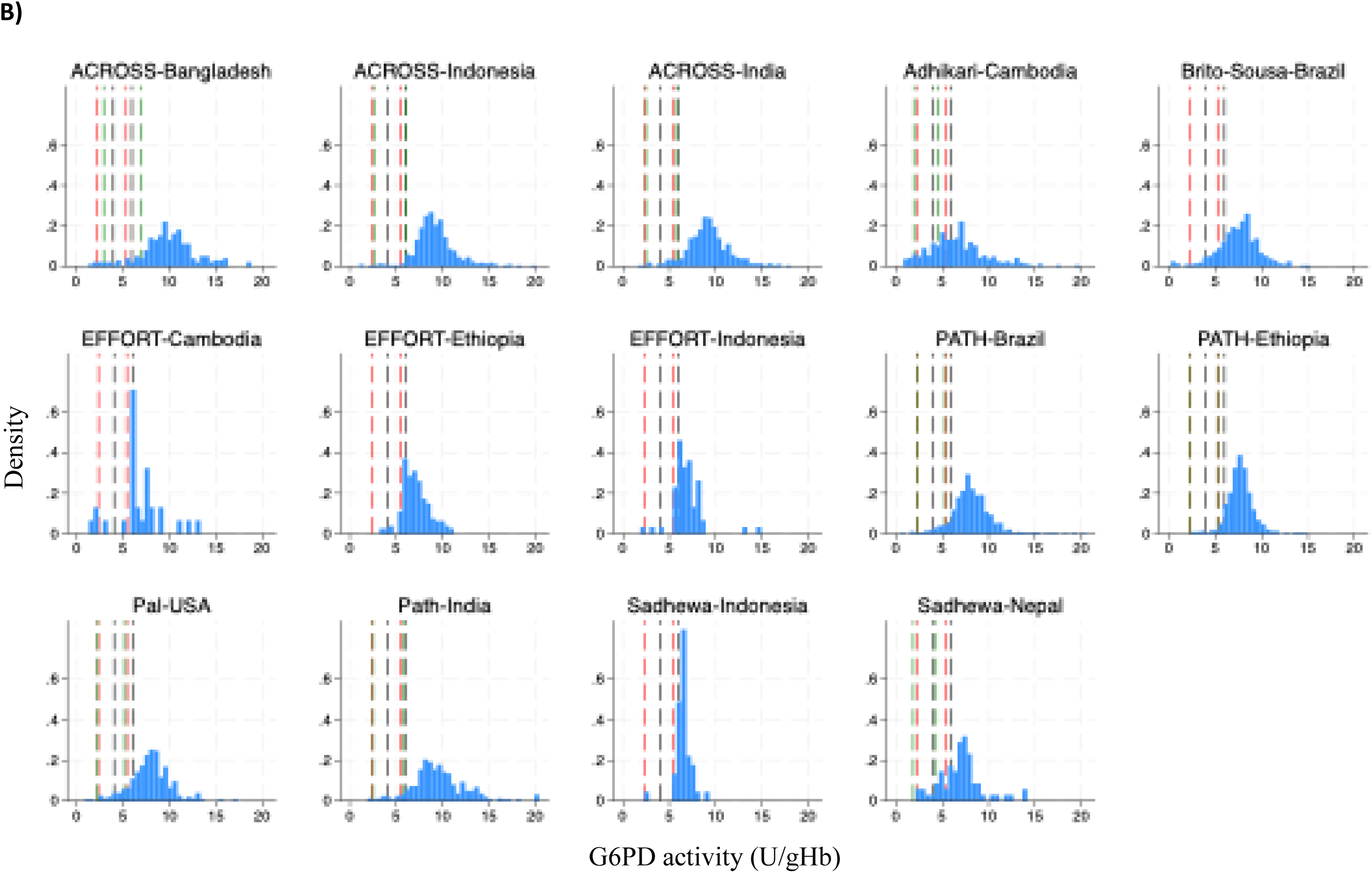
Density of G6PD activity in A) males and B) females by setting. Red dotted lines correspond to 30% and 70% activity of the universal AMM. Black dotted lines correspond to the manufacturer recommended cutoff of 4.0 U/gHb and 6.0 U/gHb. Green dotted lines correspond to 30% and 70% activity of the setting-specific AMM. Setting-specific AMMs were not available for EFFORT in Cambodia, Ethiopia and Indonesia, Sadhewa et al in Indonesia [34] and Brito-Sousa et al in Brazil [31].

**Table 1.** Demographics and baseline measures by setting.

| Country | Study | Participants | Age | Sex |  | Malaria present |  | Blood sampling method |  | Haemoglobin (g/dL) |
| --- | --- | --- | --- | --- | --- | --- | --- | --- | --- | --- |
|  |  |  |  | Male | Female | No or unknown | Yes | Venous | Capillary |  |
| Overall |  | 9,724 | 30.0 (21.0-43.0) | 5,487 (56.4%) | 4,237 (43.6%) | 7,442 (76.5%) | 2,282 (23.5%) | 786 (8.1%) | 8,938 (91.9%) | 13.5 (12.0-15.0) |
| Bangladesh | ACROSS 2019 [33] | 468 | 35.0 (15.0-45.0) | 269 (57.5%) | 199 (42.5%) | 468 (100.0%) | 0 (0.0%) | 468 (100.0%) | 0 (0.0%) | 14.4 (13.0-15.8) |
| Brazil | Brito-Sousa 2022 [31] | 1,097 | 23.0 (12.0-39.0) | 674 (61.4%) | 423 (38.6%) | 0 (0.0%) | 1,097 (100.0%) | 0 (0.0%) | 1,097 (100.0%) | 12.9 (11.3-14.4) |
|  | PATH 2019 [33] | 1,730 | 37.0 (27.0-47.0) | 785 (45.4%) | 945 (54.6%) | 1,480 (85.5%) | 250 (14.5%) | 8 (0.5%) | 1,722 (99.5%) | 13.2 (12.0-14.5) |
| Cambodia | Adhikari 2022 [32] | 1,327 | 31.0 (23.0-42.0) | 993 (74.8%) | 334 (25.2%) | 1,259 (94.9%) | 68 (5.1%) | 0 (0.0%) | 1,327 (100.0%) | 14.0 (12.4-15.7) |
|  | EFFORT 2023 | 282 | 26.0 (21.0-35.0) | 251 (89.0%) | 31 (11.0%) | 5 (1.8%) | 277 (98.2%) | 0 (0.0%) | 282 (100.0%) | 14.6 (13.3-15.7) |
| Ethiopia | EFFORT 2023 | 373 | 23.0 (19.0-30.0) | 211 (56.6%) | 162 (43.4%) | 0 (0.0%) | 373 (100.0%) | 0 (0.0%) | 373 (100.0%) | 14.0 (12.5-15.5) |
|  | PATH 2019 [33] | 1,013 | 24.0 (21.0-31.0) | 538 (53.1%) | 475 (46.9%) | 1,013 (100.0%) | 0 (0.0%) | 0 (0.0%) | 1,013 (100.0%) | 14.6 (13.2-16.2) |
| Indonesia | ACROSS 2018 | 752 | 31.0 (11.0-47.5) | 310 (41.2%) | 442 (58.8%) | 743 (98.8%) | 9 (1.2%) | 0 (0.0%) | 752 (100.0%) | 13.5 (12.4-14.9) |
|  | EFFORT 2023 | 195 | 28.0 (19.0-40.0) | 134 (68.7%) | 61 (31.3%) | 6 (3.1%) | 189 (96.9%) | 0 (0.0%) | 195 (100.0%) | 14.1 (12.6-15.4) |
|  | Sadhewa 2024 [34] | 59 | 35.0 (29.0-42.0) | 14 (23.7%) | 45 (76.3%) | 59 (100.0%) | 0 (0.0%) | 0 (0.0%) | 59 (100.0%) | 16.0 (14.6-17.1) |
| India | ACROSS 2020 [33] | 778 | 33.0 (20.0-47.0) | 359 (46.1%) | 419 (53.9%) | 759 (97.6%) | 19 (2.4%) | 250 (32.1%) | 528 (67.9%) | 13.2 (11.7-14.8) |
|  | PATH 2020 [33] | 909 | 35.0 (23.0-50.0) | 585 (64.4%) | 324 (35.6%) | 909 (100.0%) | 0 (0.0%) | 0 (0.0%) | 909 (100.0%) | 12.3 (10.8-13.8) |
| Nepal | Sadhewa 2024 [34] | 120 | 32.0 (21.0-50.0) | 51 (42.5%) | 69 (57.5%) | 120 (100.0%) | 0 (0.0%) | 60 (50.0%) | 60 (50.0%) | 14.2 (13.1-16.2) |
| USA | Pal 2021 [30] | 621 | 34.0 (26.0-48.0) | 313 (50.4%) | 308 (49.6%) | 621 (100.0%) | 0 (0.0%) | 0 (0.0%) | 621 (100.0%) | 12.4 (11.4-13.7) |

IQR – interquartile range

**Table 2.** Adjusted male medians and cutoffs by setting.

| Country | Study | Participants | 100% AMM,<br>U/gHb | 30% AMM,<br>U/gHb | Percentage (n) identified as <30% cutoff by: |  |  | 70% AMM,<br>U/gHb | Percentage identified as <70% cutoff by: |  |  |
| --- | --- | --- | --- | --- | --- | --- | --- | --- | --- | --- | --- |
|  |  |  |  |  | Manufacturer (n) | Universal (n) | Setting (n) |  | Manufacturer (n) | Universal (n) | Setting (n) |
| Overall |  | 9,724 | 7.7 | 2.3 | 7.2% (699) | 4.2% (411) | 3.9% (301) | 5.4 | 18.3% (1,777) | 11.9% (1,156) | 10.8% (830) |
| Bangladesh | ACROSS 2019 [33] | 468 | 9.9 | 3.0 | 7.7% (36) | 6.0% (28) | 7.1% (33) | 6.9 | 9.6% (45) | 8.8% (41) | 11.5% (54) |
| Brazil | Brito-Sousa 2022 [31] | 1,097 | - | - | 5.9% (65) | 2.6% (29) | - | - | 19.1% (209) | 11.9% (130) | - |
|  | PATH 2019 [33] | 1,730 | 7.5 | 2.3 | 5.5% (95) | 3.3% (57) | 3.1% (53) | 5.3 | 15.5% (268) | 9.9% (171) | 9.3% (161) |
| Cambodia | Adhikari 2022 [32] | 1,327 | 6.4 | 1.9 | 21.5% (285) | 13.3% (176) | 9.9% (132) | 4.5 | 44.1% (585) | 33.8% (449) | 24.8% (329) |
|  | EFFORT 2023 | 282 | - | - | 11.3% (32) | 10.3% (29) | - | - | 33.7% (95) | 16.7% (47) | - |
| Ethiopia | EFFORT 2023 | 373 | - | - | 2.1% (8) | 0.3% (1) | - | - | 25.2% (94) | 5.1% (19) | - |
|  | PATH 2019 [33] | 1,013 | 7.5 | 2.3 | 1.9% (19) | 0.8% (8) | 0.8% (8) | 5.3 | 7.9% (80) | 3.7% (37) | 3.7% (37) |
| Indonesia | ACROSS 2018 | 752 | 8.6 | 2.6 | 2.0% (15) | 0.9% (7) | 0.9% (7) | 6.0 | 3.9% (29) | 2.3% (17) | 3.9% (29) |
|  | EFFORT 2023 | 195 | - | - | 4.6% (9) | 2.1% (4) | - | - | 30.8% (60) | 9.2% (18) | - |
|  | Sadhewa 2024 [34] | 59 | - | - | 1.7% (1) | 1.7% (1) | - | - | 28.8% (17) | 8.5% (5) | - |
| India | ACROSS 2020 [33] | 778 | 8.3 | 2.5 | 2.3% (18) | 1.3% (10) | 1.3% (10) | 5.8 | 7.1% (55) | 4.9% (38) | 6.6% (51) |
|  | PATH 2020 [33] | 909 | 8.3 | 2.5 | 5.1% (46) | 2.9% (26) | 2.9% (26) | 5.8 | 8.6% (78) | 6.7% (61) | 7.9% (72) |
| Nepal | Sadhewa 2024 [34] | 120 | 6.2 | 1.9 | 7.5% (9) | 1.7% (2) | 0.8% (1) | 4.3 | 34.2% (41) | 25.8% (31) | 10.0% (12) |
| USA | Pal 2021 [30] | 621 | 7.3 | 2.2 | 9.8% (61) | 5.3% (33) | 5.0% (31) | 5.1 | 19.5% (121) | 14.8% (92) | 13.7% (85) |

AMM – adjusted male median; An AMM was not calculated for Sadhewa et al in Indonesia [34] due to too few male participants. An AMM was not calculated for EFFORT in Cambodia, Ethiopia and Indonesia, and Brito-Sousa et al in Brazil [31] as included participants were predominantly parasitaemic.

Two settings sampled more than 10 individuals whose G6PD activity was measured from both capillary and venous samples [33, 34]. After adjusting for age, sex, baseline Hb, and malaria status with clustering by setting, there was no difference in mean G6PD activity between capillary or venous sampling (mean difference: 0.28 U/gHb, 95% CI -1.27, 1.83).

Data were available from 6 settings on 5,064 participants with and without malaria [33, 34]. After adjusting for confounders, there was no difference in mean G6PD activity between participants with or without malaria (-0.64 U/gHb, 95% CI -1.91, 0.64). Similarly, removing parasitaemic participants did not change the percentages of participants classified as having <30% or <70% G6PD activity substantially (Table S2).

There was a non-linear association between age and G6PD activity and age and haemoglobin (Figure S1). In an exploratory analysis, measured G6PD activity decreased sharply with increasing age in children aged 1 to 5 years and plateaued thereafter, after adjusting for sex, baseline haemoglobin and blood sampling method with clustering by setting (Figure 5). This relationship was apparent in subgroups of male and female individuals. In sensitivity analyses, the age differences remained apparent when restricted to males with a haemoglobin >10 g/dL and males with G6PD activity >4.0 U/g Hb.

**Figure 5.**
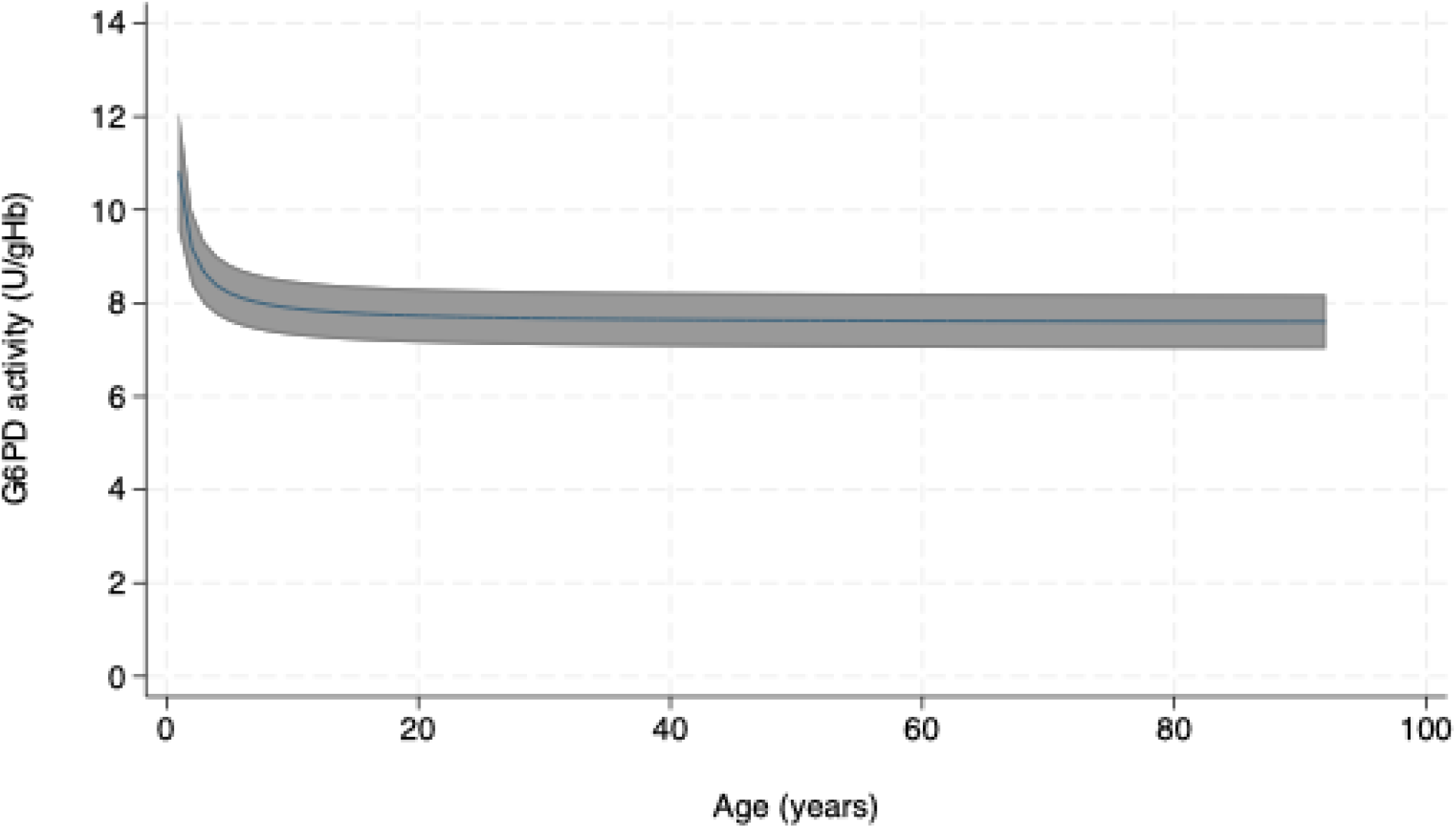
Relationship between age and G6PD activity in all individuals Based on a generalised estimating equation model with fractional polynomial terms for age, adjusted for sex, haemoglobin and blood sampling method with clustering by setting after exclusion of participants with haemoglobin <7 g/dL or >20 g/dL.

## Discussion

A reliable and accurate point-of-care diagnostic for G6PD activity is an important tool for the safe prescription of 8-aminoquinoline antimalarial drugs in vivax-endemic countries. Our individual patient data pooled meta-analysis of over 9,700 participants from 11 studies and eight countries demonstrates substantial heterogeneity in the definition of 100% activity between settings. The current manufacturer-recommended cutoffs to define deficient and intermediate G6PD activity are conservative, placing a higher proportion of individuals in the intermediate or deficient category than when applying locally calculated cutoffs. Definitions for 100% activity based on setting-specific AMMs ranged from 6.2 U/gHb to 9.9 U/gHb between settings and this was reflected by significant variation in the respective 30% and 70% cutoffs. A previous study demonstrated that, under ideal laboratory conditions, with trained laboratory staff, the Biosensor performed comparably across different settings [35]. The differences observed in our analysis could therefore be related to differences in user proficiency or inherent differences between sites.

The universal definition of 100% activity in our pooled sample was 7.7 U/gHb according to the Biosensor, and the corresponding cutoff for 30% activity (2.3 U/gHb) that was derived from this AMM appeared to exclude a substantial proportion of presumed deficient males from the G6PD deficient group based on the distribution of G6PD activities in the male population (Figure 3 and Figure 4). In comparison, across all settings most male participants at the lower end of the G6PD activity distributions were included in the deficient group using the manufacturer-recommended cutoff of 4.0 U/gHb (Figure 3 and Figure 4).

A cutoff at 30% activity defined based on spectrophotometry measurements captures the majority of male deficient patients [15]. In contrast, the Biosensor manufacturer-recommended cutoff of 4.0 U/gHb corresponds to 52% activity as determined by the universal AMM. This difference suggests there is an imperfect correlation between G6PD activity measured by spectrophotometry and G6PD activity measured by Biosensor, such that the correlation is not 1:1. Most qualitative G6PD diagnostics distinguish deficient and normal individuals at the 30% activity level, developed based on results from spectrophotometry, and have effectively guided radical cure in males over several decades [36]. Additional investigation of the relationship between Biosensor and spectrophotometric G6PD activity is warranted including assessment of haemolytic risks at various cutoffs to identify whether adjustments to the manufacturer-recommended cutoff of 4.0 U/gHb to identify G6PD deficient individuals is needed.

The manufacturer recommended cutoff of 6.0 U/gHb corresponds to 78% activity using the universal AMM and thus places a higher proportion of participants in the category of intermediate activity. Treatment with high dose primaquine radical cure (1 mg/kg/day for 7 days) potentially has a greater risk of haemolysis in heterozygous females with intermediate activity [37, 38]. A recent clinical trial in Bangladesh, Ethiopia and Indonesia provided this high dose primaquine radical cure regimen to individuals with ≥70% activity based on the local AMM. At one site the corresponding cut-off was 5 U/gHb, well below the manufacturer recommended 6.0 U/gHb cutoff for intermediate patients, with no treatment-related serious adverse events observed, and no patients developing severe anaemia (haemoglobin <5 g/dL) [39]. This study and previous studies suggest that patients with activities below 6.0 U/gHb may tolerate high dose primaquine and the optimal cutoff to determine heterozygous female patients at risk of severe haemolysis requires further investigation [40].

Neither the source of blood sampling nor malaria status impacted phenotypic G6PD activity, but G6PD activity did vary with age. Measured G6PD activity decreased from ∼11 U/gHb to ∼8 U/gHb as age increased from one to five years before plateauing thereafter. A decrease in G6PD activity over the first 6 to 12 months of life had been described previously, however, to our knowledge, this has not been observed in older children [41, 42]. This observation warrants further investigation since it may relate to either a true variation in enzyme activity, other influential parameters such as haemoglobin varying over age, or specific features associated with point of care testing with the Biosensor. If confirmed, this will have important clinical relevance, including the potential for reduced susceptibility to drug-induced haemolysis in young children and hence increased confidence in administering the radical cure in children [43]. If this finding is specific for the Biosensor, it may require an age-based adjustment in results in young children to ensure children are not falsely classified as G6PD normal. Our study has several limitations. The proportion of individuals with G6PD deficiency is known to vary substantially between and within countries [44]. Although data from nearly 10,000 individuals from eight countries were included, the study population is not necessarily representative of all populations in vivax endemic regions. Furthermore, setting-specific cutoffs may have been influenced by differences in participant age as well as sample handling, though G6PD activity remains stable for at least seven days if samples are stored between 4°C to 8°C [45, 46]. Data were not available on pregnancy, recent surgery, recent blood loss, or recent blood transfusion, preventing any adjustment for their effect on G6PD activity [8, 47, 48]. The pooled G6PD activity measurements did not have a paired spectrophotometric reference measurement, preventing assessment of systematic bias in specific settings. Due to the absence of a reference method, it was also not possible to compare the performance of universal and manufacturer recommended cutoffs against a gold-standard method, instead we had to rely on visual inspection of activity distributions.

In the absence of clinical data on haemolysis it was not possible to correlate Biosensor G6PD activity with haemolytic risks, preventing an understanding of how different cutoffs would perform in a clinical population undergoing 8-aminoquinoline therapy.

In conclusion, there was a substantial difference between the manufacturer-recommended cutoffs for G6PD deficient and intermediate individuals and those derived using the current dataset based on 30% and 70% thresholds with the G6PD Standard definitions being more conservative. Further clinical studies are needed to determine the relationship between spectrophotometry and Biosensor G6PD activity measurements and the cutoffs that best predict the risk of drug-induced haemolysis.

## Data Availability

The data supporting this meta-analysis are available upon request from the individual investigators who provided the data for analysis

## Acknowledgements

We thank all participants and staff who were involved in these clinical trials at all the sites. This research has been supported by the Australian Government through the Partnerships for a Healthy Region (PHR) initiative. The views expressed in this publication are the author’s alone and are not necessarily the views of the Australian Government.

## Data sharing

Study data can be obtained by request to the corresponding authors of the included studies.

---

**Checklist S1.** PRISMA-IPD Checklist of items to include when reporting a systematic review and meta-analysis of individual participant data (IPD)

| PRISMA-IPD<br>Section/topic | Item<br>No | Checklist item | Reported<br>on page |
| --- | --- | --- | --- |
| Title |  |  |  |
| Title | 1 | Identify the report as a systematic review and meta-analysis of individual participant data. | 1 |
| Abstract |  |  |  |
| Structured<br>summary | 2 | Provide a structured summary including as applicable: | 2 |
|  |  | **Background:** state research question and main objectives, with information on participants, interventions, comparators and outcomes. |  |
|  |  | **Methods:** report eligibility criteria; data sources including dates of last bibliographic search or elicitation, noting that IPD were sought; methods of assessing risk of bias. |  |
|  |  | **Results:** provide number and type of studies and participants identified and number (%) obtained; summary effect estimates for main outcomes (benefits and harms) with confidence intervals and measures of statistical heterogeneity. Describe the direction and size of summary effects in terms meaningful to those who would put findings into practice. |  |
|  |  | **Discussion:** state main strengths and limitations of the evidence, general interpretation of the results and any important implications. |  |
|  |  | **Other:** report primary funding source, registration number and registry name for the systematic review and IPD meta-analysis. |  |
| Introduction |  |  |  |
| Rationale | 3 | Describe the rationale for the review in the context of what is already known. | 3-4 |
| Objectives | 4 | Provide an explicit statement of the questions being addressed with reference, as applicable, to participants, interventions, comparisons, outcomes and study design (PICOS). Include any hypotheses that relate to particular types of participant-level subgroups. | 4 |
| Methods |  |  |  |
| Protocol and registration | 5 | Indicate if a protocol exists and where it can be accessed. If available, provide registration information including registration number and registry name. Provide publication details, if applicable. | 5 |
| Eligibility criteria | 6 | Specify inclusion and exclusion criteria including those relating to participants, interventions, comparisons, outcomes, study design and characteristics (e.g. years when conducted, required minimum follow-up). Note whether these were applied at the study or individual level i.e. whether eligible participants were included (and ineligible participants excluded) from a study that included a wider population than specified by the review inclusion criteria. The rationale for criteria should be stated. | 5 |
| Identifying studies - | 7 | Describe all methods of identifying published and unpublished studies including, as applicable: which bibliographic databases were searched with dates of coverage; details of any hand searching including of conference proceedings; use of study registers | 5 |
| information sources |  | and agency or company databases; contact with the original research team and experts in the field; open adverts and surveys. Give the date of last search or elicitation. |  |
| Identifying studies - search | 8 | Present the full electronic search strategy for at least one database, including any limits used, such that it could be repeated. | 5 |
| Study selection processes | 9 | State the process for determining which studies were eligible for inclusion. | 5 |
| Data collection processes | 10 | Describe how IPD were requested, collected and managed, including any processes for querying and confirming data with investigators. If IPD were not sought from any eligible study, the reason for this should be stated (for each such study). | 5 |
|  |  | If applicable, describe how any studies for which IPD were not available were dealt with. This should include whether, how and what aggregate data were sought or extracted from study reports and publications (such as extracting data independently in duplicate) and any processes for obtaining and confirming these data with investigators. |  |
| Data items | 11 | Describe how the information and variables to be collected were chosen. List and define all study level and participant level data that were sought, including baseline and follow-up information. If applicable, describe methods of standardising or translating variables within the IPD datasets to ensure common scales or measurements across studies. | 5 |
| IPD integrity | A1 | Describe what aspects of IPD were subject to data checking (such as sequence generation, data consistency and completeness, baseline imbalance) and how this was done. | 5 |
| Risk of bias assessment in individual studies. | 12 | Describe methods used to assess risk of bias in the individual studies and whether this was applied separately for each outcome. If applicable, describe how findings of IPD checking were used to inform the assessment. Report if and how risk of bias assessment was used in any data synthesis. | 6 |
| Specification of outcomes and effect measures | 13 | State all treatment comparisons of interests. State all outcomes addressed and define them in detail. State whether they were pre-specified for the review and, if applicable, whether they were primary/main or secondary/additional outcomes. Give the principal measures of effect (such as risk ratio, hazard ratio, difference in means) used for each outcome. | 6 |
| Synthesis methods | 14 | Describe the meta-analysis methods used to synthesise IPD. Specify any statistical methods and models used. Issues should include (but are not restricted to):<br>• Use of a one-stage or two-stage approach.<br>• How effect estimates were generated separately within each study and combined across studies (where applicable).<br>• Specification of one-stage models (where applicable) including how clustering of patients within studies was accounted for.<br>• Use of fixed or random effects models and any other model assumptions, such as proportional hazards.<br>• How (summary) survival curves were generated (where applicable).<br>• Methods for quantifying statistical heterogeneity (such as $I^2$ and $t^2$).<br>• How studies providing IPD and not providing IPD were analysed together (where applicable). | 6 |
|  |  | How missing data within the IPD were dealt with (where applicable). |  |
| Exploration of variation in effects | A2 | If applicable, describe any methods used to explore variation in effects by study or participant level characteristics (such as estimation of interactions between effect and covariates). State all participant-level characteristics that were analysed as potential effect modifiers, and whether these were pre-specified. | 6 |
| Risk of bias across studies | 15 | Specify any assessment of risk of bias relating to the accumulated body of evidence, including any pertaining to not obtaining IPD for particular studies, outcomes or other variables. | N/A |
| Additional analyses | 16 | Describe methods of any additional analyses, including sensitivity analyses. State which of these were pre-specified. | 6 |
| **Results** |  |  |  |
| Study selection and IPD obtained | 17 | Give numbers of studies screened, assessed for eligibility, and included in the systematic review with reasons for exclusions at each stage. Indicate the number of studies and participants for which IPD were sought and for which IPD were obtained. For those studies where IPD were not available, give the numbers of studies and participants for which aggregate data were available. Report reasons for non-availability of IPD. Include a flow diagram. | 6, Figure 1 |
| Study characteristics | 18 | For each study, present information on key study and participant characteristics (such as description of interventions, numbers of participants, demographic data, unavailability of outcomes, funding source, and if applicable duration of follow-up). Provide (main) citations for each study. Where applicable, also report similar study characteristics for any studies not providing IPD. | Table 1 |
| IPD integrity | A3 | Report any important issues identified in checking IPD or state that there were none. | Nil |
| Risk of bias within studies | 19 | Present data on risk of bias assessments. If applicable, describe whether data checking led to the up-weighting or down-weighting of these assessments. Consider how any potential bias impacts on the robustness of meta-analysis conclusions. | Table S1 |
| Results of individual studies | 20 | For each comparison and for each main outcome (benefit or harm), for each individual study report the number of eligible participants for which data were obtained and show simple summary data for each intervention group (including, where applicable, the number of events), effect estimates and confidence intervals. These may be tabulated or included on a forest plot. | 7 |
| Results of syntheses | 21 | Present summary effects for each meta-analysis undertaken, including confidence intervals and measures of statistical heterogeneity. State whether the analysis was pre-specified, and report the numbers of studies and participants and, where applicable, the number of events on which it is based. | 7 |
|  |  | When exploring variation in effects due to patient or study characteristics, present summary interaction estimates for each characteristic examined, including confidence intervals and measures of statistical heterogeneity. State whether the analysis was pre-specified. State whether any interaction is consistent across trials. |  |
|  |  | Provide a description of the direction and size of effect in terms meaningful to those who would put findings into practice. |  |
| Risk of bias across studies | 22 | Present results of any assessment of risk of bias relating to the accumulated body of evidence, including any pertaining to the availability and representativeness of available studies, outcomes or other variables. | N/A |
| Additional analyses | 23 | Give results of any additional analyses (e.g. sensitivity analyses). If applicable, this should also include any analyses that incorporate aggregate data for studies that do not have IPD. If applicable, summarise the main meta-analysis results following the inclusion or exclusion of studies for which IPD were not available. | 7-8 |
| **Discussion** |  |  |  |
| Summary of evidence | 24 | Summarise the main findings, including the strength of evidence for each main outcome. | 8-10 |
| Strengths and limitations | 25 | Discuss any important strengths and limitations of the evidence including the benefits of access to IPD and any limitations arising from IPD that were not available. | 8-10 |
| Conclusions | 26 | Provide a general interpretation of the findings in the context of other evidence. | 8-10 |
| Implications | A4 | Consider relevance to key groups (such as policy makers, service providers and service users). Consider implications for future research. | 9-10 |
| **Funding** |  |  |  |
| Funding | 27 | Describe sources of funding and other support (such as supply of IPD), and the role in the systematic review of those providing such support. | 10 |

© Reproduced with permission of the PRISMA IPD Group, which encourages sharing and reuse for non-commercial purposes

Table S1. QUADAS analysis

**Table S2.** Percentage of participants identified by cutoffs by setting after exclusion of participants with malaria.

| Country | Study | Participants | Percentage (n) identified as <30% cutoff by: |  |  | Percentage (n) identified as <70% cutoff by: |  |  |
| --- | --- | --- | --- | --- | --- | --- | --- | --- |
|  |  |  | Manufacturer (n) | Universal (n) | Setting (n) | Manufacturer (n) | Universal (n) | Setting (n) |
| Overall |  | N=7,442 | 7.5% (557) | 4.4% (329) | 3.8% (281) | 16.6% (1,232) | 11.9% (888) | 10.6% (782) |
| Bangladesh | ACROSS 2019 [33] | N=468 | 7.7% (36) | 6.0% (28) | 7.1% (33) | 9.6% (45) | 8.8% (41) | 11.5% (54) |
| Brazil | PATH 2019 [33] | N=1,480 | 5.3% (79) | 3.1% (46) | 2.8% (42) | 13.9% (206) | 9.2% (136) | 8.6% (128) |
| Cambodia | Adhikari 2022 [32] | N=1,259 | 21.6% (272) | 13.3% (167) | 9.8% (124) | 44.2% (556) | 34.0% (428) | 25.0% (315) |
|  | EFFORT 2023 | N=5 | 40.0% (2) | 40.0% (2) | - | 40.0% (2) | 40.0% (2) | - |
| Ethiopia | PATH 2019 [33] | N=1,013 | 1.9% (19) | 0.8% (8) | 0.8% (8) | 7.9% (80) | 3.7% (37) | 3.7% (37) |
| Indonesia | ACROSS 2018 | N=743 | 2.0% (15) | 0.9% (7) | 0.9% (7) | 3.9% (29) | 2.3% (17) | 3.9% (29) |
|  | EFFORT 2023 | N=6 | 0.0% (0) | 0.0% (0) | - | 50.0% (3) | 16.7% (1) | - |
|  | Sadhewa 2024 [34] | N=59 | 1.7% (1) | 1.7% (1) | - | 28.8% (17) | 8.5% (5) | - |
| India | ACROSS 2020 [33] | N=759 | 2.2% (17) | 1.2% (9) | 1.2% (9) | 7.1% (54) | 4.9% (37) | 6.6% (50) |
|  | PATH 2020 [33] | N=909 | 5.1% (46) | 2.9% (26) | 2.9% (26) | 8.6% (78) | 6.7% (61) | 7.9% (72) |
| Nepal | Sadhewa 2024 [34] | N=120 | 7.5% (9) | 1.7% (2) | 0.8% (1) | 34.2% (41) | 25.8% (31) | 10.0% (12) |
| USA | Pal 2021 [30] | N=621 | 9.8% (61) | 5.3% (33) | 5.0% (31) | 19.5% (121) | 14.8% (92) | 13.7% (85) |

4 A setting specific cutoff was not available for EFFORT in Cambodia and Indonesia and Sadhewa et al in Indonesia [34] due to too few male patients to 5 estimate an adjusted male median.

**Figure S1.**
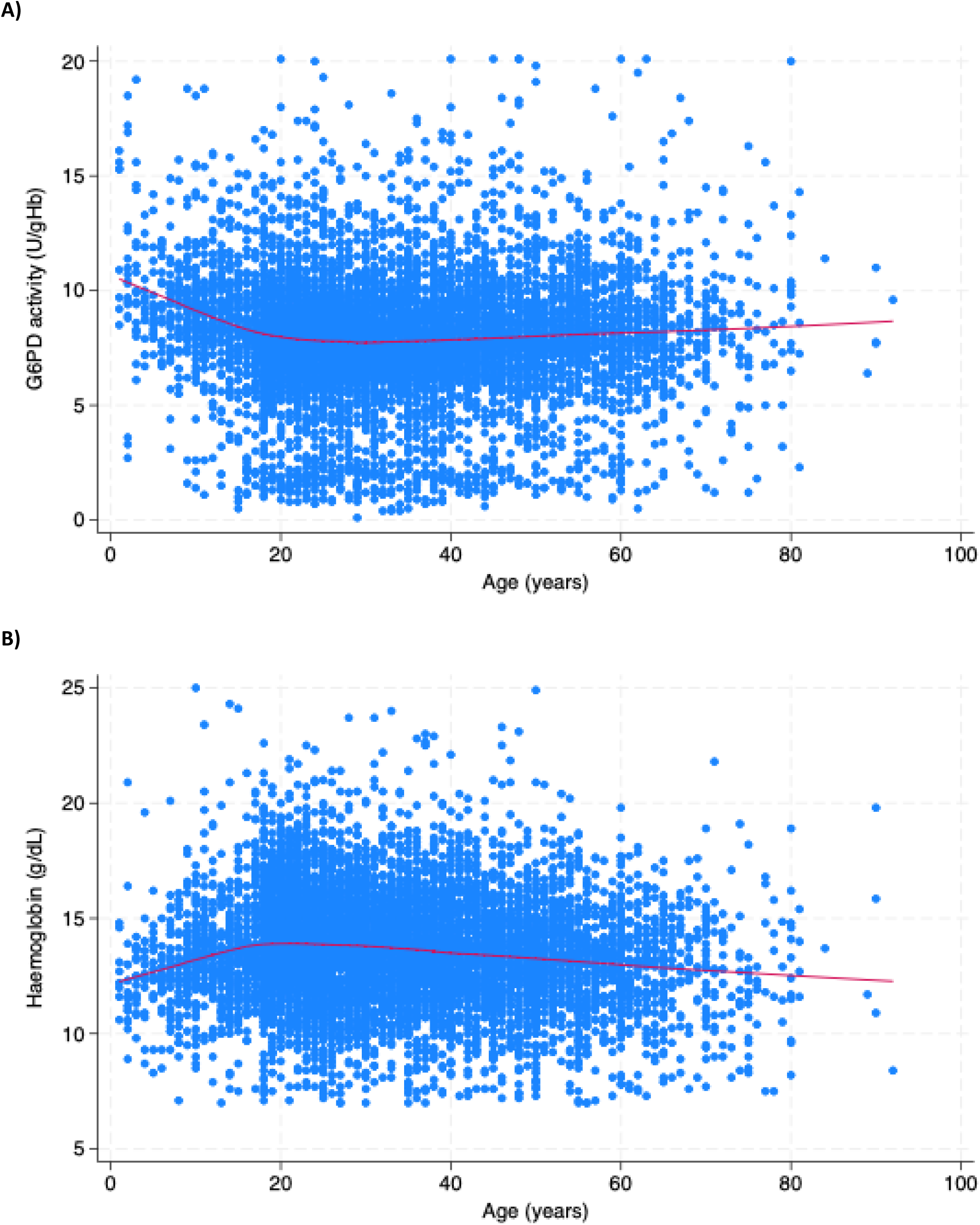
Scatter plot of A) G6PD activity (U/gHb) versus age and B) haemoglobin (g/dL) versus age A) A lowess curve using Cleveland’s tricube weighting function with a bandwidth of 0.8 is fitted to both figures. Red line = smoothed trend

